# Comparative Analysis of Machine Learning Models for Efficient Low Back Pain Prediction Using Demographic and Lifestyle Factors

**DOI:** 10.1101/2023.10.29.23297737

**Authors:** Jun-hee Kim

**Affiliations:** Yeonsedae-gil, Maeji-ri, Heungeop-myeon, Wonju-si, Gangwon-do, 26493, Laboratory of KEMA AI Research (KAIR), Department of Physical Therapy, College of Software and Digital Healthcare Convergence, Yonsei University, Wonju, South Korea

## Abstract

**Background:** Low back pain (LBP) is one of the most frequently occurring musculoskeletal disorders, and factors such as lifestyle as well as individual characteristics are associated with LBP. The purpose of this study was to develop and compare efficient low back pain prediction models using easily obtainable demographic and lifestyle factors.

**Methods:** Data from adult men and women aged 50 years or older collected from the Korean National Health and Nutrition Examination Survey (KNHANES) were used. The dataset included 22 predictor variables, including demographic, physical activity, occupational, and lifestyle factors. Four machine learning algorithms, including XGBoost, LGBM, CatBoost, and RandomForest, were used to develop predictive models.

**Results:** All models achieved an accuracy greater than 0.8, with the LGBM model outperforming the others with an accuracy of 0.830. The CatBoost model had the highest sensitivity (0.804), while the LGBM model showed the highest specificity (0.884) and F1-Score (0.821). Feature importance analysis revealed that EQ-5D was the most critical variable across all models.

**Conclusion:** In this study, an efficient LBP prediction model was developed using easily accessible variables. Using this model, it may be helpful to identify the risk of LBP in advance or establish prevention strategies in subjects who have difficulty accessing medical facilities.

## Introduction

Low back pain (LBP) is one of the most common musculoskeletal disorders experienced by adult men and women in most countries (1). It is estimated that up to 80% of the population will experience LBP at least once in their lifetime (2). LBP is prevalent and a leading cause of years lived with disability (YLD) (1). The Global Burden of Disease (GBD), Injuries and Risk Factors study estimated the age-standardized rate of YLD due to LBP to be 832 per 100,000 people, and the number of people suffering from LBP worldwide is expected to increase from 619 million in 2020 to 843 million in 2050 (1). The reported socioeconomic cost of LBP in the United States, estimates ranged from US$18.5 billion to US$28.2 billion (3).

LBP has been shown to be complexly related to uncontrollable factors such as age and gender and controllable factors such as obesity, physical activity, and lifestyle patterns (1,4). In the GBD study, the incidence rate of LBP in women was 19% higher than that in men, and LBP increased with age, reaching a peak between the ages of 80 and 89 (1,5). In particular, it has been reported that in the case of elderly people, the severity of pain becomes more severe as age increases, and the likelihood of developing chronic LBP is high (6). These changes are reported to occur due to physical changes due to aging (spinal degeneration, spinal disease, physical inactivity) (6). Controllable factors contributing to 38.8% of YLD due to LBP included occupational factors, smoking, and BMI (1). Occupation-related physical activities such as vibration, lifting, bending, and twisting have been reported as potential risk factors for LBP (7). Current smokers reported a higher incidence of LBP in the past month and were particularly associated with chronic LBP and severe LBP (8). People who smoked but quit smoking had a higher prevalence of LBP than those who had never smoked, but it was lower than those who were currently smoking (8). Overweight and obesity have been reported to increase the incidence and severity of LBP in the past 12 months (9).

Machine learning is a technology in which a computer analyzes data consisting of various variables and learns patterns of relationships between them to build a predictive model based on a large amount of data (10). Recently, research has been actively conducted to develop machine learning models based on variables related to LBP, such as an individual’s physical, lifestyle, and environmental factors (11,12). In these studies, machine learning models for LBP were developed to predict the presence or absence of LBP, analyze risk factors, and predict the patient’s pain level or prognosis (11– 14). In addition, a machine learning model that predicts LBP by analyzing changes in brain structure with computer vision technology that processes images or a model that predicts the classification of LBP high-risk and low-risk groups by analyzing changes in specific movement patterns with sensors capable of measuring three-dimensional motions was developed, and a model that predicts LBP by interpreting medical reports on diagnostic images with natural language processing technology was even developed (15–18).

Variable selection and prediction performance are important factors that determine the efficiency and usefulness of a machine learning model (10). Correct variable selection and optimization helps improve model accuracy and reduce unnecessary complexity, which is very important for prediction and decision-making using machine learning (10). From this perspective, the ease of acquiring variables will also be an important factor in building efficient and practical machine learning models (10,19). In other words, it is important to build an efficient model to increase prediction performance while being easy to acquire variables and incurring low cost (10,19). However, high-performance machine learning models for LBP have used not easy-to-acquire variables that require medical diagnostic equipment and expert measurement, whereas LBP models made with simple measurement variables have a problem of poor predictive performance (11,15,16,18). Studies that built LBP prediction models by analyzing data collected in the form of magnetic resonance imaging (MRI), computed tomography (CT), and 3D images using computer vision technology all showed high accuracy of 0.8 or higher (15). However, in a study by Lamichhane et al. (2021), the accuracy was shown to be 0.745, when a predictive model was made of morphological changes in cerebral cortical thickness (CT) and resting-state functional connectivity (rsFC) as a potential brain biomarker for LBP (18). Also, Shim et al. (2021) studied a risk factor prediction model for chronic LBP using patient demographic variables, comorbidities variables, psychological variables, lifestyle variables, health indicator variables, but a total of 26 predictor variables were used in building the final model of the study and the accuracy was relatively low at 0.717 (11).

Therefore, the purpose of this study was as follows. First, the goal was to develop an efficient LBP prediction model that could achieve high performance based on variables that are easy to obtain. In developing a high-performance model, demographic and lifestyle factors were used as variables that were easy to measure, and various algorithms were used to develop models and compare their performance. The second was to analyze which variables might be related as risk factors for predicting LBP. The importance of variables in the optimized models was analyzed to determine which variables were most important in predicting LBP.

## Methods

### Data Collection

The flowchart of this study for the development of LBP prediction machine learning model is shown in Figure 1. In this study, datasets from the 5th and 6th Korean National Health and Nutrition Survey (KNHANES) conducted by the Korea Centers for Disease Control and Prevention from 2010 to 2015 were used. The subjects of this study were all over 50 years of age, and 4,228 people who had LBP for more than 30 days within the last 3 months were assigned to the LBP group, and 13,987 were assigned to the painless control group. In the dataset, a total of 22 predictor variables were selected, including demographic variables such as age, gender, height, and weight, physical activity variables such as vigorous physical activity, strength training, and walking days, occupational variables such as occupation type and average working hours, and lifestyle variables such as smoking, housing type, and health-related quality of life (Table 1).

**Table 1.**
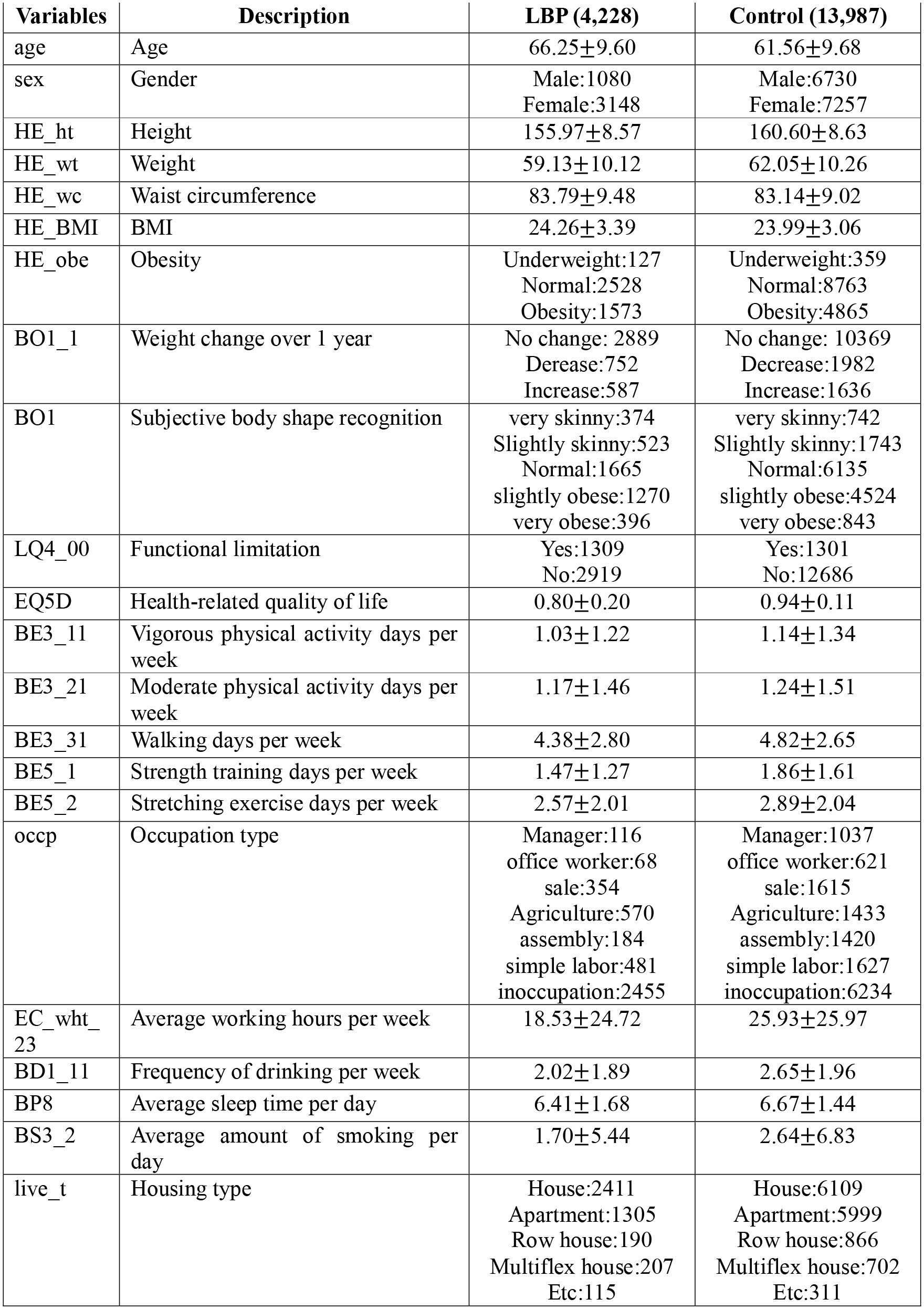
Description of variables.

**Figure 1.**
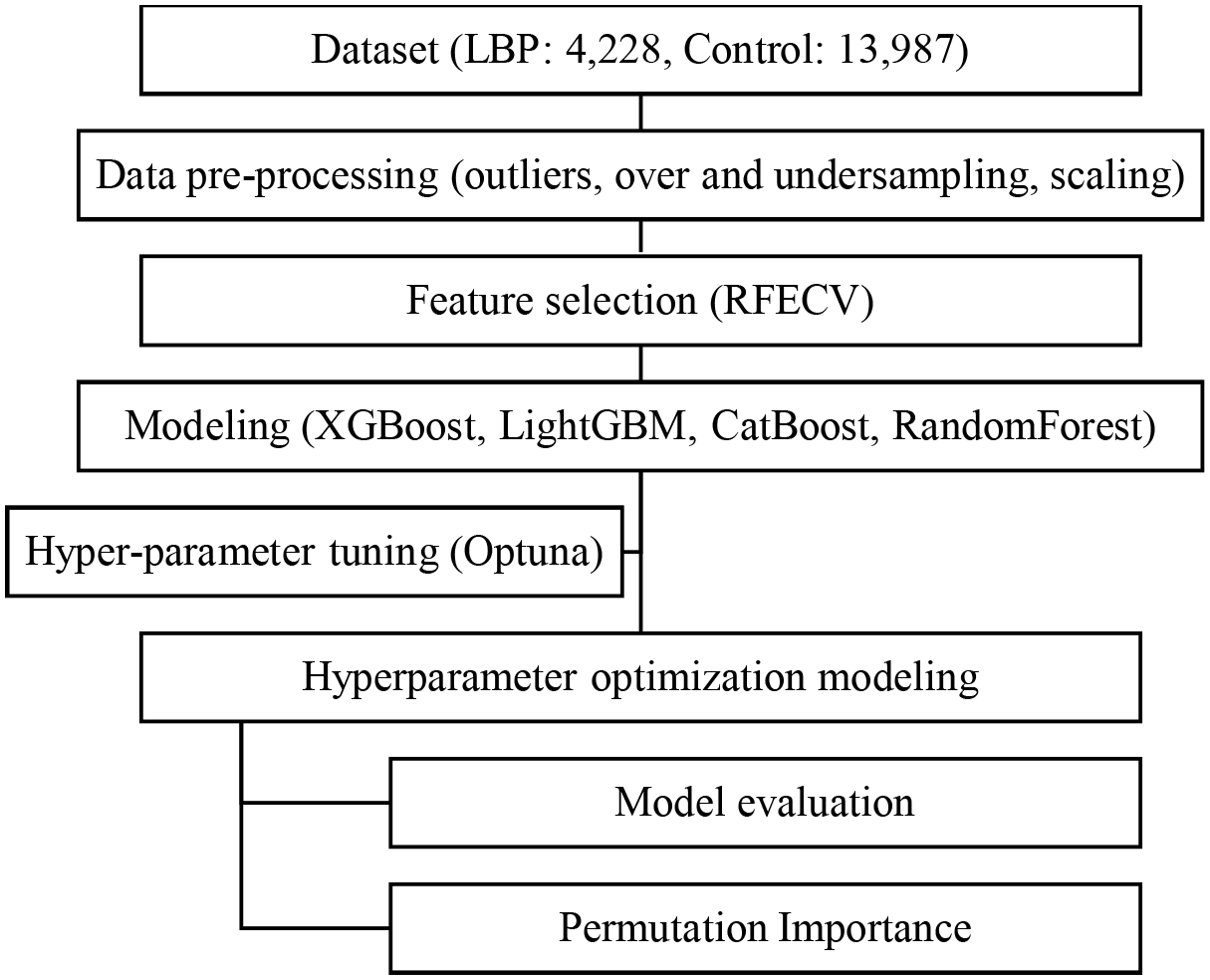
Flowchart of the study.

### Data Pre-processing

To process the imbalance data between groups, this study applied RandomUnderSampler, which randomly reduces the size of the dataset, and synthetic minority oversampling technique (SMOTE), which randomly increases the size of the dataset, and matched the ratio between groups to 1:1. Outliers were detected and processed by the IQR (Interquartile Range) method. A value less than Q1 - 1.5 * IQR was converted to that value, and a value greater than Q3 + 1.5 * IQR was also converted to that value. Numerical variables were scaled with StandardScaler to make the mean zero and the standard deviation one. Categorical variables were converted to LabelEncoder function. After the pre-processing process, the data were divided into training and test sets at a 9:1 ratio for model training and performance evaluation.

### Feature selection

Recursive Feature Elimination with Cross-Validation (RFECV) was used as a feature selection technique (20). RFECV is used to remove unnecessary variables that degrade the performance of the model. RFECV finds the optimal feature set for each model through an iterative process (20).

### Model Training and Hyper-parameter Tuning

In this study, each model was developed using three boosting algorithms and one bagging algorithm. The algorithms used were XGBoost, which provides fast learning speed and high accuracy; LGBM, which provides the fastest processing speed for large datasets; CatBoost, which shows high performance in categorical data processing; and RandomForest, which combines the results of multiple decision trees. Models were constructed with variables selected by each algorithm and RFECV technique (Table 2). The models for each algorithm were tuned using the Optuna library, a tool for hyperparameter optimization. Through this, each model automatically searches for optimal hyperparameters and adjusts them to maximize model performance. After adjusting the hyperparameters using Optuna, the final models were trained with Stratified K-fold cross-validation (CV) using training set data to set the CV to 5.

**Table 2.** RFECV Variable Selection and Scores.

| **Model** | **Selected variables** | **Score** |
| --- | --- | --- |
| XGB | 'sex', 'live_t', 'LQ4_00', 'EQ5D', 'BO1_1', 'BE3_21' | 0.827 |
| LGBM | ALL | 0.828 |
| CatBoost | ALL | 0.828 |
| Random Forest | ALL | 0.830 |

### Model evaluation

The evaluation of model performance used test set data that was not used in the model training process to prevent overfitting and objectively evaluate performance. The evaluation of the model was performed based on the Confusion Matrix of the test set for each model. These evaluations include Accuracy, Sensitivity, Specificity, and F1-Score. The performance of each model was comprehensively evaluated and compared through the indicators. The feature importance for each trained model was evaluated by permutation importance, which relatively reflected the effect of each variable on the performance of the model.

## Results

### Feature selection

The variables selected in the model for each algorithm and the accuracy when training the model with those variables are presented in Table 2. Compared to other algorithms, the model of the XGBoost algorithm showed the highest training accuracy when constructing a model with the fewest six variables. All other algorithm models showed the highest training accuracy when constructing a model using all 22 variables.

### Model performance

The hyperparameters and training scores of each algorithm optimized using the Optuna library are shown in Table 3. The confusion matrix of each model using the test set is shown in Figure 2, and the evaluation performances are shown in Table 4. The overall accuracy of the LGBM model of 0.830 was the highest. In addition, the LGBM model had the highest specificity and F-1 score of 0.884 and 0.821, respectively. However, the CatBoost model with a sensitivity of 0.804 was the highest. The RandomForest model showed the lowest accuracy and specificity of 0.818 and 0.834 respectively, while the XGBoost model showed the lowest sensitivity and F-1 score of 0.775 and 0.812, respectively.

**Table 3.**
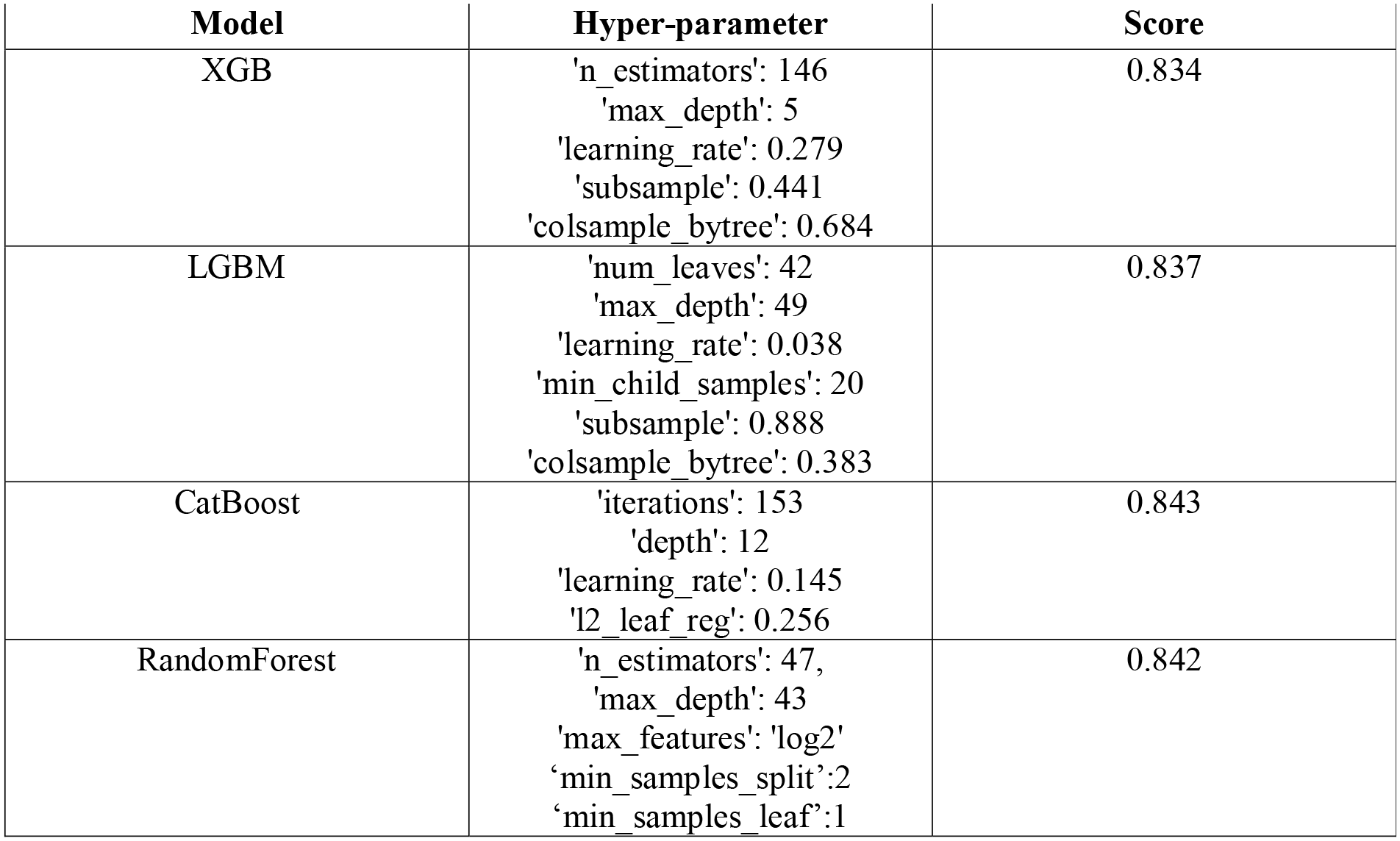
Optuna Model Hyperparameters and Scores.

**Table 4.** Test data performance of optimized models.

| Model | Accuracy | Sensitivity | Specificity | F-1 Score |
| --- | --- | --- | --- | --- |
| XGB | 0.821 | 0.775 | 0.866 | 0.812 |
| LGBM | 0.830 | 0.777 | 0.884 | 0.821 |
| Random Forest | 0.818 | 0.801 | 0.834 | 0.815 |
| CatBoost | 0.824 | 0.804 | 0.845 | 0.821 |

**Figure 2.**
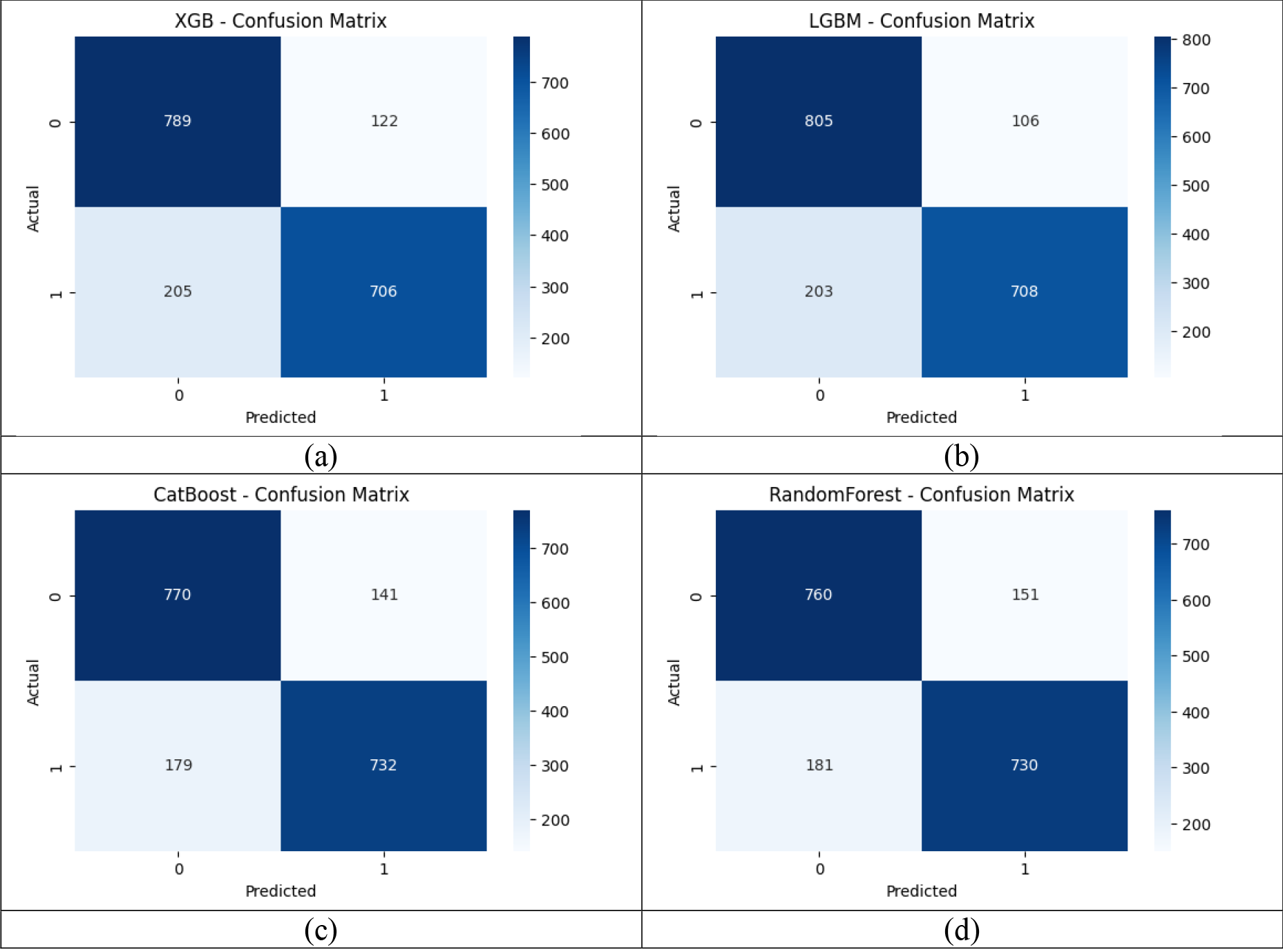
Confusion matrix: XGBoost (a), LGBM (b), CatBoost (c), RandomForest (d)

### Feature importance

The importance of each variable in each model is presented in Figure 3. In all models, the most important variable in the composition of the models was EQ-5D. In the RandomForest and LGBM model, demographic data such as age, height, weight, BMI, and waist circumference were highly ranked as important variables in constructing the model. In contrast, the Catboost model showed relatively high variable importance for lifestyle pattern variables such as sleep time, housing type, frequency of drinking, walking days per week, and stretching days per week.

**Figure 3.**
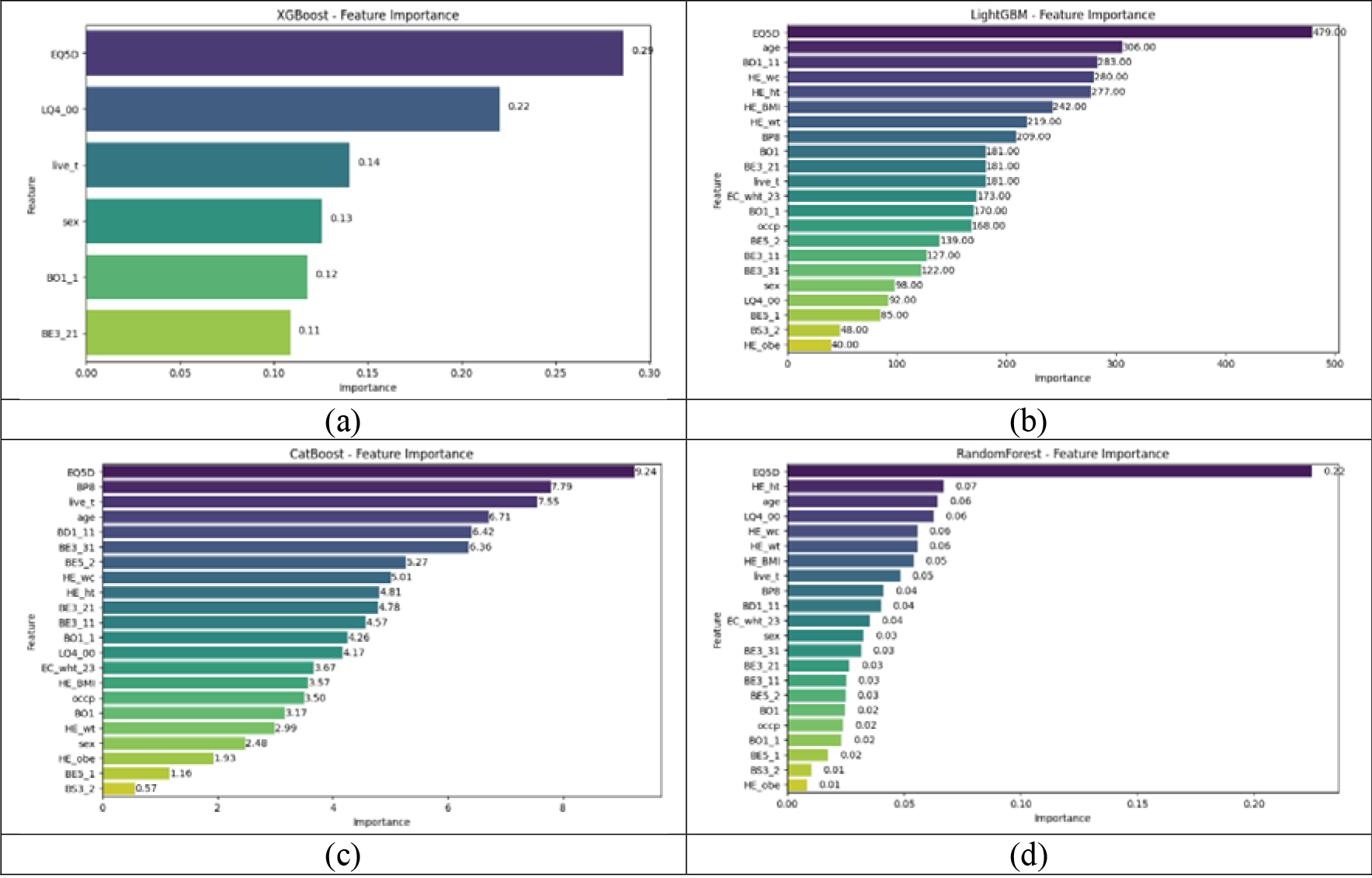
Feature importance: XGBoost (a), LGBM (b), CatBoost (c), RandomForest (d)

## Discussion

The purpose of this study was to develop an efficient LBP prediction model with high performance based on easily acquired demographic, physical-related, and lifestyle pattern data. In addition, it was to identify variables related to LBP through the feature importance in the predictive models. LBP prediction models were developed by applying various boosting and bagging algorithms and technologies for optimizing performance. In constructing each optimized model, a minimum of 6 to a maximum of 22 variables were used as variables. The accuracy of the models developed in this study all exceeded 0.8. In terms of overall performance, accuracy, the LGBM model was the best, and in terms of sensitivity, the CatBoost model was the best. The most important variable in constructing all models was EQ-5D.

Prior to this study, Shim et al. (2021) developed and compared a machine learning model to predict LBP, which used 26 variables, including comorbid diseases, psychological factors, and health indicator variables, as well as demographic and lifestyle variables, and the performance of machine learning models ranged from 0.656 to 0.716 (11). In this study, unlike their model, a model was developed in which the number of variables was reduced to 6 or 22, excluding variables that were difficult to obtain, which should be measured by medical diagnostic equipment or measured by experts, and the performance of the model was 0.818 to 0.830. The model of this study could have been improved over the model of the previous study based on the following reasons. First, the difference in performance would have occurred due to the difference in the amount of data. In the previous study, the model was developed with data of 6,119 people over two years, and in this study, the model was developed with data of 18,215 people over six years. Larger datasets are more likely to reflect the representativeness of the subjects (21). The more data is used to develop the model, the more likely it is not overfitting, but the lack of data can lead to overfitting and inaccurate generalization (21). In addition, complex patterns and interrelationships may appear more clearly on larger datasets (21). The second is probably due to the difference in the number and selection of variables. As variables increase, so does the complexity of the model. Using complex models for higher-dimensional data can make the model unstable and reduce the ability to generalize (22). Increasing dimensions as the number of variables increases creates a curse of dimensions where space becomes larger and rarer, and more data is needed to develop the model (22). In addition, high correlations between variables can lead to multicollinearity, which makes the model’s stability and interpretation difficult (22). When it comes to the characteristics of variable, the model’s performance can be degraded if important variables are not selected, or unnecessary variables are used in the model (22). In this study, a smaller number of variables were used to develop the model, and RFECV techniques were applied to control the number of variables to prevent the exclusion of important variables or the inclusion of unnecessary variables in the model. Third, it is probably due to an imbalance between the number of LBP data and the number of non-LBP data. If the number of samples in one data is significantly smaller than that of the others, the model will be skewed to data with more samples, resulting in insufficient learning of patterns in minority data and limited predictive performance (23). The model in this study, developed with medical data where the data imbalance problem occurs primarily, solved the data imbalance with techniques such as SMOTE and RandomUnderSampler to avoid the above and problems. For these reasons, the model in this study would have performed better compared to the previous study.

Nevertheless, in addition to this study, models with higher performance in lumbar-related diseases or LBP predictions have been developed. When using variables acquired by imaging equipment such as MRI or 3D scanning devices that require expert measurement rather than the type of variables used in the predictive model developed in this study, it was possible to create a model for predicting lumbar spine-related diseases that showed excellent performance or to further improve the performance of the model for predicting LBP (24–26). Adankon et al. (2012) developed a model to classify the type of scoliosis with a support vector machine algorithm based on data acquired by a non-invasive 3D scan method, and the classification accuracy of the model was 0.95 (24). Based on MRI data of the lumbar spine, Ruiz et al. (2015) developed a model that extracts disk shape features and detects contour abnormalities with the Gradient Vector Flow algorithm, and the model showed accuracy of 0.90 or more (25). Ketola et al. (2021) developed a model to predict LBP by applying texture analysis to the two lowest lumbar discs (L4-L5 and L5-S1) on T2-weighted MRI (26). The accuracy of the developed model was 0.83, the specificity was 0.83, and the sensitivity was 0.82 (26). Compared with the model of this study, the accuracy was similar, and the specificity was slightly lower, but the sensitivity was slightly higher. As a result of the above studies, models developed with variables measured by medical diagnostic equipment or experts showed stable high performance. It is meaningful that these models elicit high performance based on a small number of variables. Nevertheless, it may be considered a disadvantage that these variables require measurement by medical diagnostic equipment or experts. The models constructed in this study do not require medical diagnostic equipment or expert skills in measuring those variables, even though many kinds of variables are required. This point will be highly utilized in predicting or managing LBP in the elderly population or people living in areas with poor access to health care.

A Model has also been developed to identify variables representing differences in texture features in MRI images between subjects with or without LBP, rather than LBP prediction models (27). This model was built not to predict LBP, but to identify variables that simply represent high feature importance (27). From this point of view, the risk factors related to LBP could be identified through the feature importance of the predictive model constructed in this study. The most important variable in the development of the model for this study was EQ-5D. EQ-5D is a tool that comprehensively represents physical activity, lifestyle patterns, and psychological factors to evaluate health-related quality of life with a total of 5 items including mobility, self-care, user activity, pain/discomfort, and anxiety/depression. (28). This highlights the relationship between the multidimensional nature of LBP and the general health and quality of life of individuals. In RandomForest and LGBM models, demographic variables such as age, height, weight, BMI, and waist circumference have been shown as very important predictors. These findings are consistent with previous studies suggesting that age and body composition may be a risk factor for LBP in relation to changes in spinal health and musculoskeletal disorders (1). On the other hand, the CatBoost model placed relative importance on lifestyle variables including sleep time, housing type, frequency of drinking, walking days per week and stretching days per week. This suggests that aspects such as sleep quality, physical activity, and living environment may affect an individual’s sensitivity to LBP. For example, improper sleep, sedentary behavior, or poor living conditions can contribute to musculoskeletal problems. Differences in varying importance between models imply that LBP predictions benefit from an integrated approach. Demographic factors provide insight into the physical aspects of LBP risk, while lifestyle patterns provide a broader perspective, covering behavioral and environmental impacts. Therefore, a comprehensive LBP prediction model should consider both health-related quality of life and various personal characteristics. These findings have practical implications for healthcare practitioners. They suggest that strategies for preventing and managing LBP should be tailored to individuals based on their unique profiles. Personalized approaches that consider health-related quality of life, demographic factors, and lifestyle choices may be more effective at reducing the burden of LBP.

The study has several limitations. First, this study is based on data from the National Health and Nutrition Survey of Korea (KNHANES), and the study subjects were limited to adults over 50 years of age. Therefore, it may be difficult to apply generalizations of results and models to populations of different age groups or countries. Second, because this study built a model using easily obtainable variables, other important variables used in diagnosing LBP or variables obtained through medical diagnostic tools were not included in the model, which may result in lower accuracy compared to other models. Third, methods such as SMOTE and RandomUnderSampler have been used to address data imbalance between the LBP and control groups, but these methods do not always yield ideal predictive results when evaluated with real-world different data. Fourth, this study considered various variables to predict LBP, but other variables such as posture and movement, which are the direct cause of LBP, or variables that interact with them should also be considered. Fifth, the algorithms used in this study are efficient at improving predictive performance, but their complexity makes it difficult to interpret the causal relationship of variables. Future studies will require the use of representative data from various age groups and countries. In addition to the variables used in this study, models with different types of data and variables can be considered for combining medical diagnostic data variables or for a deep understanding of the causes of LBP. This could improve the predictive accuracy of the model and help to better understand the causes of future LBP. In addition to the sampling techniques currently used, models could be developed as more effective sampling techniques to address medical data imbalances. Finally, future research should be developed as an algorithm that can reduce the complexity of the model and better interpret causal relationships between variables. This will be more intuitive and descriptive for healthcare practitioners and patients to understand and trust model results.

## Conclusion

In this study, an efficient LBP prediction model with an accuracy of over 0.8 was developed through various boosting and bagging algorithms utilizing easily measurable demographic, physical, and lifestyle variables. The performance of these models showed that the EQ-5D variable, which encompasses physical activity, lifestyle, and psychological factors, was the most important variable in all models, reflecting the multifaceted nature of LBP. This study provides an efficient machine learning model development approach as an adjunctive tool for LBP prediction and management and may provide valuable insights to healthcare practitioners aiming to develop personalized strategies to alleviate the burden of LBP.

## Data Availability

All data produced are available online at: the Korea Centers for Disease Control and Prevention

https://knhanes.kdca.go.kr/knhanes/main.do

